# A Community-Based Integrated Partnership Approach to Living Well with Dementia: Evaluating the Impact of the Sage House Model

**DOI:** 10.64898/2026.01.10.26343830

**Authors:** Rachel. L King, Stuart Warren, Elizabeth Vass, Benjamin. T Sharpe, Kian Beaumont, Stewart Seymour, Sam Bell, Antonina Pereira

**Author notes:** Correspondence Dr Rachel King, University of Chichester, Bishop Otter Campus, College Lane, Chichester, West Sussex, PO19 6PE.

## Abstract

A key societal priority is to support people with dementia and their care partners to live well, particularly through improved access to high-quality information, continuity of care, and consistent post-diagnostic support. The Sage House Model represents a novel, community-based approach to integrated dementia care, co-locating diagnostic assessment, tailored support services, psychosocial interventions, and opportunities for social engagement within a single accessible centre. By reducing fragmentation across health, social care, and voluntary sector provision, the model is designed to address key determinants of living well with dementia. The present study examined whether the Sage House Model is associated with key wellbeing outcomes for both people with dementia and care partners. Using a natural experiment design, outcomes were compared between participants accessing the Sage House centre in Tangmere and those receiving standard care. The sample included 135 people with dementia (*M*_age_ = 74.64, *SD* = 8.30) and 129 care partners (*M*_age_ = 67.23, *SD* = 9.84). A series of ANCOVAs indicated that people with dementia accessing Sage House reported significantly higher quality of life (*p* = .004, ω^2^ = .06), life satisfaction (*p* = .004, ω^2^ = .07), and wellbeing (*p* = .044, ω^2^ = .03) compared with those receiving standard care. In addition, care partners with access to Sage House reported significantly greater needs-based QoL, particularly in relation to improved access to information and support (*p* = .005, ω^2^ = .07). These findings provide initial empirical support for community-based integrated dementia care in enhancing wellbeing for people with dementia and their care partners, and highlight the value of co-located, multicomponent dementia services in addressing post-diagnostic gaps in real-world care.

---

In the UK it is estimated that there are currently 982,000 people with dementia, with projections indicating a rise to between 1.4 and 1.7 million by 2040 (Alzheimer’s Society, 2024; Chen et al., 2023). Globally, approximately 57 million people were living with dementia in 2021, with a predicted 10 million new cases per year (WHO, 2025). Dementia has a profound impact on quality of life, functional independence, and wellbeing for both those diagnosed and their care partners (Agüero-Torres et al., 1998; Banerjee et al., 2006; Gale et al., 2018; Giebel et al., 2014; Gilsenan et al., 2023; Nakayama et al., 2017; Reid & O’Brien, 2021). The increasing prevalence of dementia, coupled with its impact on both those diagnosed and those providing care, underscores the need to optimise interventions and support systems that enhance wellbeing. In the present paper, we will evaluate a novel model of integrated dementia support, known as the *Sage House Model*.

## The Sage House Model

The Sage House Model of integrated dementia support is a community-based, multicomponent supportive care approach designed as a comprehensive “one-stop” hub for people with dementia and their care partners. Through a collaborative community partnership strategy, it integrates NHS diagnostic services, psychosocial interventions, and practical support within a single dementia-friendly centre, bringing together health, social, and voluntary sector provision across the care pathway. Consistent with definitions of multicomponent supportive care (Guzzon et al., 2023), the model integrates multiple non-pharmacological strategies, including psychosocial interventions and care partner support, within a single service framework. Refined through co-design to reflect community needs, the model leverages local connections to deliver flexible, personalised care and is inclusive from pre-diagnosis through to end-of-life, supporting continuity. This structure allows the model to be evaluated not as a single intervention, but as an integrated system of support operating under routine service conditions. Developed by the charity Dementia Support, the model has been operationalised in Sage House, a dedicated centre located in Tangmere, UK, since May 2018 ([Sage House Model | Dementia Support](https://nap.dementiasupport.org.uk/sage-house-model)), aligning with current policy priorities for continuous, accessible, community-based integrated care (Department of Health and Social Care, 2025).

### Components of the Sage House Model

The Sage House Model is comprised of several core components, including on-site diagnostic assessments, allocation of a dedicated support worker (Wayfinder) and provision of respite daycare (Daybreaks). There is also a range of optional psychosocial interventions enabling personalised engagement that aligns with individual preferences and evolving needs throughout the dementia trajectory. These psychosocial interventions include cognitive stimulation, physical activity, social and educational programmes, and enrichment activities. Practical support is also embedded within the model, with integrated access to services such as legal advice, personal care, and information about locally provided resources. The scope of the components integrated within a single, purpose-built centre is a distinguishing and significant feature of this Sage House model.

When considering the scope of provision provided, it is helpful to consider the comprehensive framework of dementia support outlined by Bamford et al. (2021). The Sage House Model provides support that spans all five thematic domains identified, including the timely identification and management of needs, understanding and managing dementia, psychological and emotional wellbeing, practical support, and the requirement for the integration of care. Of the 20 components underpinning these themes, the Sage House Model directly addresses 14 and contributes supportively to the remaining six, which are clinical in focus (e.g., diagnosis, care planning). This framework has been proposed as a benchmark to assist commissioners in identifying effective provision of local services (Bamford et al., 2023). In this context, the Sage House Model is particularly notable for its scope and ability to integrate existing local community services collaboratively. See Table 1 for an overview.

**Table 1.** Overview of Components of the Sage House Model and their Alignment with Bamford et al. (2021) Framework for Post-Diagnostic Dementia Support

| Type | Component | Description | Bamford et al., 2021 Themes/Components |
| --- | --- | --- | --- |
| Diagnostic Support | *Dementia Assessment Team | NHS Dementia Assessment Service (DAS) provides diagnostic meeting and ongoing review on site. | Timely identification and management of needs. |
| Information, Support and Guidance | *Wayfinding | A single point of contact who provide practical and emotional support from pre-diagnosis to end of life. Accessible via phone or in-person appointments. | Cross-cutting theme of integrating support, as well as multiple components in understanding and managing dementia, psychological & emotional wellbeing and practical support |
|  | Training | Training courses to support understanding of dementia, support carers and to train local businesses to provide dementia-friendly services. E.g., Empowering Carers Course and Dementia Supporters. | Information provision, developing strategies for living with dementia and managing behavioural symptoms of dementia |
|  | Solicitor & Citizen Advice | Local solicitor and citizens advice team provide support on dementia related legal and practical matters provided onsite. | Information provision, advocacy and support with finances. |
| Psychosocial Intervention | Cognition | Activities that promote cognitive health. E.g., Cognitive Stimulation and Reminiscence Therapy. | Improving/ maintaining cognition, meaningful activities, supporting emotional wellbeing, peer support and supporting relationships. |
|  | Physical | Activities that promote physical health. E.g., Yoga, Chair Aerobics and Carpet Bowls. |  |
|  | Social | Activities that promote social wellbeing. E.g., Daisy Café, Chatter Tables and Support Groups for both people with dementia and care partners. |  |
|  | Enrichment | Activities that promote wellbeing. E.g., Singing, Painting, Events, and Knit & Natter |  |
|  | Outreach | Activities and peer support groups delivered within community settings. |  |
| Care | Personal Care | Onsite provision of bespoke room fitted with accessibility bath/ shower facilities, onsite hairdressing facilities with local dementia-friendly hairdresser and other professionals that offer services on site such as footcare and therapies (e.g., massage) | Practical support for example having a break for the care partners and psychological and emotional support. |
|  | *Respite | On site 6-hour daybreaks service with personal care support included. Food provided from the café and meaningful activities engaged in whilst at Daybreaks. |  |

Note: Asterisk indicates core components.

### Evaluating the Impact of the Sage House Model

While the Sage House Model offers a broad range of interventions, it is important to evaluate its effectiveness in delivering meaningful societal outcomes. Enhancing dementia support has long been a key societal objective, reflected in both UK and global policy for over a decade (DoH, 2009, 2011, 2016a; NHS England, 2016; WHO, 2017). This objective is underpinned by the concept of “living well with dementia”, which looks to achieve the best possible health across physical, mental, and social dimensions (Institute of Medicine, 2015). Notably, this concept acknowledges the challenges of living with dementia while striving to optimise QoL within those constraints. Living well encompasses quality of life, subjective wellbeing, and life satisfaction, reflecting individuals’ perceptions of meaning, purpose, and emotional balance (Martyr et al., 2018; Quinn et al., 2022; Tournier et al., 2023; van Horik et al., 2022). These domains are consistently identified as priorities by people living with dementia and their care partners (Quinn et al., 2022; Tournier et al., 2023). Therefore, “living well” with dementia provides a relevant, person-centred framework to assess the Sage House Model’s impact.

Another key societal objective is promoting care partner wellbeing (Department of Health, 2009, 2011, 2016a, 2016b; NHS England, 2016; World Health Organization, 2017). Research has identified factors influencing care partner QoL, including psychological and physical health, the quality of the caregiver relationship and social networks (Clare et al., 2019; Farina et al., 2017). It is also important to note that care partners often face barriers that affect their QoL and caregiving capacity, such as limited access to information, training, and support services (Arblaster & Brennen, 2022; Hargreaves, 2022; MacLeod et al., 2017; Peterson et al., 2016). The care dyad relationship is bidirectional with carer stress, isolation and competence also influencing the person with dementia’s ability to “live well” (Clare et al., 2019; Quinn et al., 2020). Consequently, a further aim of the current study is to evaluate whether attending the Sage House Centre in Tangmere can improve QoL for carers due to improved access to information and support.

### Study Aim and Research Question

This study aims to assess whether the Sage House model improves wellbeing outcomes. Specifically, it will address the research question: does access to the Sage House centre in Tangmere influence key wellbeing outcomes for people living with dementia and their care partners? The results from this study are intended not only as an evaluation at the individual level, but also as a consideration of how integrated community services can address fragmentation and variability in post-diagnostic dementia care.

## Method

### Ethical Approval

This study was approved by the University of Chichester Ethics Committee (2223_05). Procedures followed the principles outlined in the declaration of Helsinki and participants provided informed consent prior to taking part.

### Participants

People with dementia and care partners were recruited via word of mouth, leaflets, and the Join Dementia Research network (https://www.joindementiaresearch.nihr.ac.uk/).

At Sage House, the Wayfinding team supported recruitment by sharing study details and facilitating contact with the research team. Data collection occurred over 18 months with a minimum sample size of 30 targeted for each group (Zhang et al., 2022).

### Participants with Dementia

Participants required a diagnosis of mild cognitive impairment or dementia, capacity to provide informed consent, and the absence of severe sensory or comprehension impairments that would impact their engagement with the study materials. 137 individuals with dementia took part, however, two withdrew resulting in a final sample of 135 (89 in the control group, 46 with access to the Sage House centre in Tangmere). See supplementary Table 1 for sample characteristics.

### Care Partners

Participants were required to be over 18 and have direct experience caring for someone with dementia. Participants were required to be the primary caregiver and be currently actively caring for an individual with dementia. 129 participants took part (72 in the control group and 57 in the Sage House group). See supplementary Table 2 for sample characteristics

### Experimental Design

A natural experiment design was adopted comparing participants with access to the Tangmere Sage House centre to a usual care comparator reflecting locally available, non-integrated services. The design and analysis were informed by the GRACE checklist for high-quality observational studies of comparative effectiveness (Dreyer et al., 2010, 2016). See Supplementary Table 3 for further details. Both the intervention and standard care control have strong clinical relevance, reflecting real-world practice with sufficient exposure to detect effects, standardised measures have been employed and collected systematically in the same period. Restricting access to the centre would not have been feasible or ethically appropriate meaning that randomisation was not available within the context of the current design.

### Intervention and Control

The Sage House Model was examined by working with participants who had unlimited access to the Sage House centre in Tangmere. For individuals with dementia, the average time they had been attending the centre was 11.95 months (*SD* = 12.53, range: 1 month to 4 years). For care partners, average attendance was 18.14 months (*SD* = 21.92, range: 2 weeks to 9 years). Within the last three months there was an average 19.98 visits (*SD* = 16.51), with a maximum of 91 visits. Engagement varied based on personal preference and need. Participants in the control group had access to standard healthcare, defined here as access to routine medical services and any locally available support services that were not delivered through a coordinated or integrated care pathway.

### Service and Stakeholder Engagement

A series of meetings were held with Sage House representatives to inform study design and guide the selection of outcome measures. Those representatives contributed dual lived experience, drawing on both personal experience as family carers and professional roles supporting people living with dementia at the Sage House centre, thereby providing broad, contextually relevant expertise. Representatives were also invited to provide feedback during manuscript development.

### Materials

Wellbeing measures were selected in consultation with Sage House representatives and guided by a review by Clarke et al. (2020) of wellbeing measures for people with dementia and prior research on “living well” with dementia (Clarke et al., 2020; Martyr et al., 2018). For care partners, comparable measures were selected alongside a needs-based QoL measure focused on access to information and support within the caregiving role (Horton et al., 2021; Oyebode et al., 2018).

## Quality of Life

### Quality of Life in Alzheimer’s Disease

(QoL-AD; Logsdon et al., 2002)

The QoL-AD is a 13-item self-report measure for assessing QoL in people with dementia across domains including energy, memory, mood, and relationships. Responses are rated on a 4-point Likert scale (1–4), yielding a total score that ranges from 13–52, with higher scores indicating better QoL. In the current study, internal consistency was good (α = .88). The QoL-AD has demonstrated strong psychometric properties (Thorgrimsen, 2003) and has been used previously in “living well” research (Lamont et al., 2020).

### Scales measuring the Impact of Dementia on Carers

*(SIDECAR;* Horton et al., 2021; Oyebode et al., 2018).

SIDECAR is a dementia-specific, needs-based QoL measure developed from interviews with care partners (Oyebode et al., 2018). It comprises three subscales: direct impact of caring (18 items, range 0–18), indirect impact of caring (10 items, range 0–10), and access to support and information (11 items, range 0–11). Higher scores indicate greater unmet need and therefore lower QoL. The SIDECAR meets COSMIN standards and has demonstrated strong psychometric properties (Horton et al., 2021). In the current sample, internal consistency was good for the direct impact subscale (α = .82) and adequate for the access to support and information subscale (α = .73). Internal consistency for the indirect impact subscale was poor (α = .58) and therefore this subscale was excluded from the analyses.

## Life Satisfaction

### Satisfaction with Life

(SwLS; Pavot & Diener, 1993, 2008)

The SWLS is a 5-item self-report measure of life satisfaction, rated on a 7-point Likert scale (1–7). Total scores range from 5–35, with a score of 20 indicating a neutral level of life satisfaction and higher scores reflecting greater satisfaction. In the present sample, internal consistency was good for both people with dementia and care partners (α = .82). The SWLS has demonstrated strong psychometric properties (Pavot et al., 1991) and has been used previously in “living well” dementia research (Lamont et al., 2020).

## Wellbeing

### World Health Organisation Wellbeing Index

(WHO-5; Topp et al., 2015; World Health Organisation, 1998)

The WHO-5 is a 5-item self-report measure of subjective wellbeing assessing experiences over the previous two weeks. Items are rated on a 6-point Likert scale (0–5), yielding raw scores from 0–25, which are multiplied by four to produce a total score ranging from 0–100, with higher scores indicating greater wellbeing. In the current sample, the internal consistency was good (α = .81). The WHO-5 has been validated for use with older adults (Heun et al., 1999) and has been used previously in “living well” dementia research (Lamont et al., 2020).

### Warwick-Edinburgh Mental Wellbeing Scale

(WEMWBS; Tennant et al., 2007)

The WEMWBS is a 14-item self-report measure of subjective wellbeing, rated on a 5-point Likert scale (1–5). Total scores range from 14–70, with higher scores indicating greater wellbeing. In the current sample, internal consistency was excellent (α = .90). The WEMWBS has demonstrated strong psychometric properties and sensitivity to change following intervention (Maheswaran et al., 2012; Tennant et al., 2007a).

## Control Measures

To assess group comparability, self/proxy reported functional capacity was measured using the Frenchay Activities Index (FAI; Schuling et al., 1993). Additionally, cognitive impairment severity was assessed in the dementia group using the Montreal Cognitive Assessment (MoCA; Nasreddine et al., 2005) and caregiver burden was measured in the care partner sample using the Caregiver Burden Inventory (CBI; Novak & Guest, 1989).

### Frenchay Activities Index

(FAI; Schuling et al., 1993).

The FAI is a 15-item self-report measure assessing engagement in instrumental activities of daily living over the past three or six months, including tasks such as meal preparation, reading, and work. Items are rated on a 4-point Likert scale (0–3), with higher scores indicating greater functional capacity. In the current study, internal consistency was good for both self-reports from participants with dementia (α = .84) and proxy reports from care partners (α = .83). The FAI has shown strong psychometric properties in older adults and stroke populations (Schuling et al., 1993) and good agreement between self- and proxy ratings (Segal & Schall, 1994).

### Montreal Cognitive Assessment

(MoCA; Nasreddine et al., 2005)

The MoCA is a brief (∼10-minute) cognitive screening measure assessing visuospatial/executive function, naming, memory, attention, language, abstraction, and orientation. Total scores range from 0–30, with scores of 26 or above indicating normal cognition (Dautzenberg et al., 2020). The MoCA is widely used for its sensitivity in detecting cognitive impairment and its applicability across diverse populations and clinical settings (Nasreddine et al., 2005).

### Caregiver Burden Inventory

(CBI; Novak & Guest, 1989)

The CBI is a 24-item self-report measure assessing caregiver burden across domains including time dependency, personal development, physical and emotional health, and social relationships. Items are rated on a 5-point Likert scale (0–4). In the current sample, internal consistency was excellent (α = .90). The CBI has demonstrated strong utility in assessing caregiver burden among those supporting people with dementia (Marvardi et al., 2005).

## Study Protocol

### Procedure

Participants were recruited via advertisements at Sage House and the Join Dementia Research Network. Interested individuals contacted the research team or approached if permission had been given and they met the inclusion criteria. Upon expressing interest, they received an information sheet and scheduled an appointment, either in person or via Teams. At the start of the session participants reviewed the information sheet, asked questions, and provided informed consent. In all sessions measures were administered via Qualtrics (Qualtrics, Provo, UT) to ensure consistency across format. A research team member guided participants through the protocol, which included three sections with natural breaks. For individuals with dementia, the sections covered 1) demographics and health economics information, 2) wellbeing measures, and 3) control measures and an open-ended question. For care partners, the sections covered 1) demographics and wellbeing, 2) impact of caring, 3) proxy measures for the person they support along with an open-ended question. Sessions typically lasted one hour, with variation based on individual need. Participants were debriefed upon completion. Results from the health economics evaluation and open question are reported elsewhere.

### Data Handling

Data handling and analysis were conducted in R version 4.3.1 (R Core Team, 2023) using packages such as tidyverse (Wickham et al, 2019), here (Müller, 2020), psych (Revelle, 2023), effectsize (Ben-Shachar, Lüdecke & Makowski, 2020), and emmeans (Lenth, 2023). Prior to analysis statistical assumptions were checked to ensure parametric assessment was appropriate and outliers beyond three standard deviations from the mean were removed and checked for impact on statistical analysis.

### Data Analysis

The level of functional independence has previously been shown to impact the ability to “live well” with dementia (Martyr et al., 2019), and in the present study it was correlated with the wellbeing dependent variables and therefore it was considered as a covariate. A series of one-way between subjects ANCOVAs were utilised to examine group differences in average performance on each wellbeing variable. As multiple wellbeing measures were examined reported P-values have been corrected using the false discovery rate (Benjamini & Hochberg, 1995). Non-significant results are reported as per current recommendations (Visentin et al., 2020).

## Results

### People with Dementia: Living Well with Dementia Outcomes

#### Quality of Life

After controlling for FAI, the group with access to Sage House had significantly higher QoL-AD scores (*M* = 37.86, 95% CI [36.10, 39.62]) as compared to the standard care control group (*M* = 34.32, 95% CI [33.08, 35.57]), *F*(1, 131) = 9.89, *p* = .004, ω^2^ = .06. A medium effect size was observed, with the group that had access to Sage House scoring on average 3.54 points higher, indicating better QoL (Figure 1A). One outlier (> 3 SD) was removed from the data; this removal did not impact the statistical conclusions.

**Figure 1.**
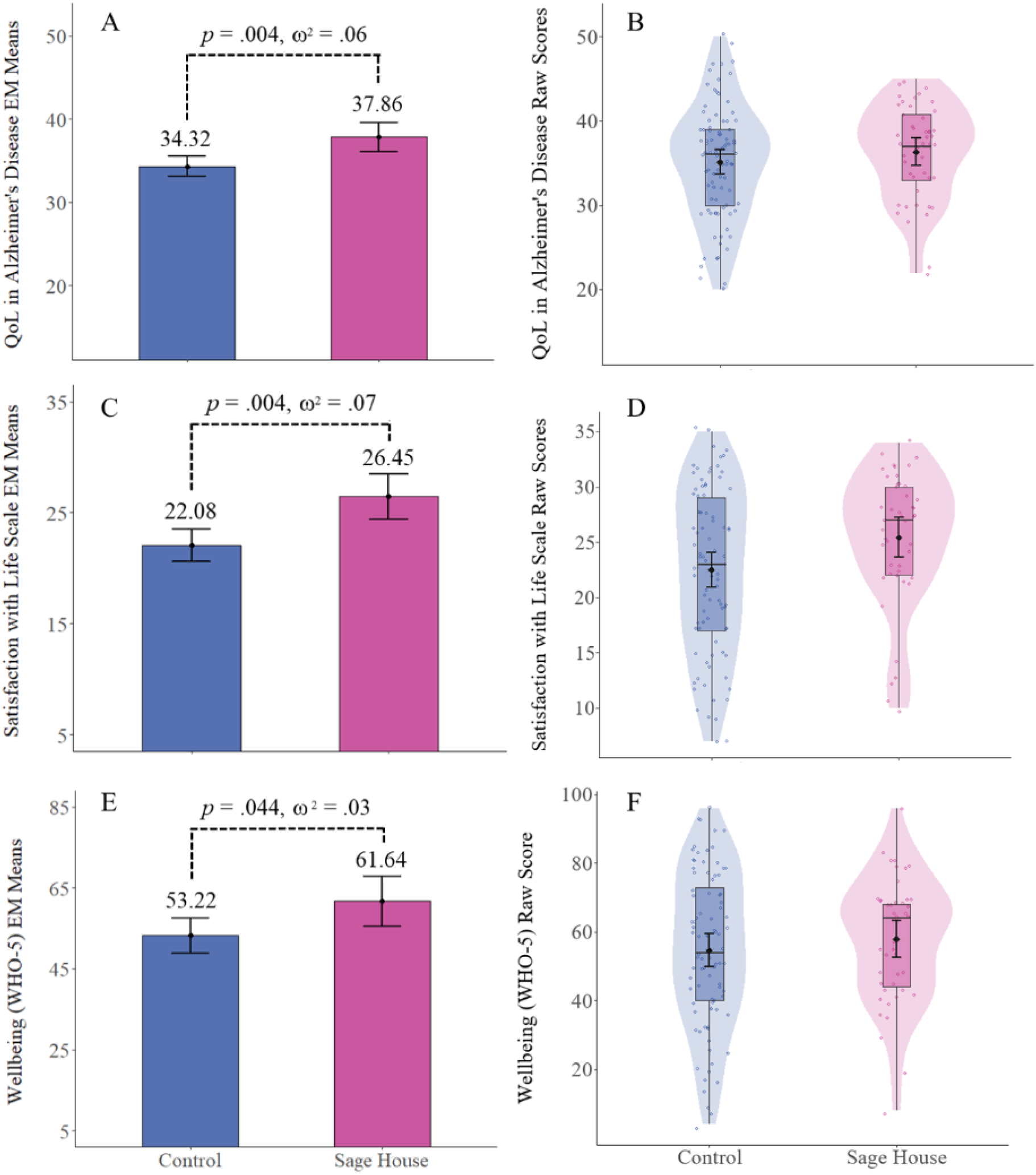
Wellbeing outcomes split by Sage House group. A & B) AD-QoL group comparison. C & D) SwLS group comparison. E & F) WHO-5 group comparison. Left hand graphs represent estimated marginal means after correcting for FAI. FDR corrected P-Values displayed. Right hand violin graphs represent raw means, 95% CI and the data distribution.

#### Life Satisfaction

After controlling for FAI, the group with access to Sage House had significantly higher SwLS scores (*M* = 26.45, 95% CI [24.39, 28.51]) as compared to the standard care control group (*M* = 22.08, 95% CI [20.63, 23.53]), *F*(1, 132) = 11.12, *p* = .004, ω^2^ = .07. A medium effect size was observed, with the group that had access to Sage House scoring on average 4.37 points higher indicating greater life satisfaction (Figure 1C).

#### Subjective Wellbeing

After controlling for FAI, the group with access to Sage House had significantly higher WHO-5 scores (*M* = 61.64, 95% CI [55.49, 67.80]) as compared to the standard care control group (*M* = 53.22, 95% CI [48.89, 57.55]), *F*(1, 132) = 4.64, *p* = .044, ω^2^ = .03. A small effect size was observed, with the group that had access to Sage House scoring on average 8.42 points higher, indicating greater subjective wellbeing (Figure 1E).

## Care Partners: Wellbeing Outcomes

### Needs-Based QoL: Impact of access to Information & Support

After controlling for FAI, the group with access to Sage House had significantly greater Needs-Based QoL related to access to support and information (*M* = 3.41, 95% CI [2.75, 4.07]) as compared the standard care control group (*M* = 4.93, 95% CI [4.34, 5.52]), *F*(1, 126) = 11.31, *p* = .005, ω^2^ = .07. A medium effect size was observed, with the group that had access to Sage House scoring on average 1.52 points lower. In this instance lower scores indicate that their need for access to support and information was being met more effectively (Figure 2A). All other comparisons were non-significant.

**Figure 2.**
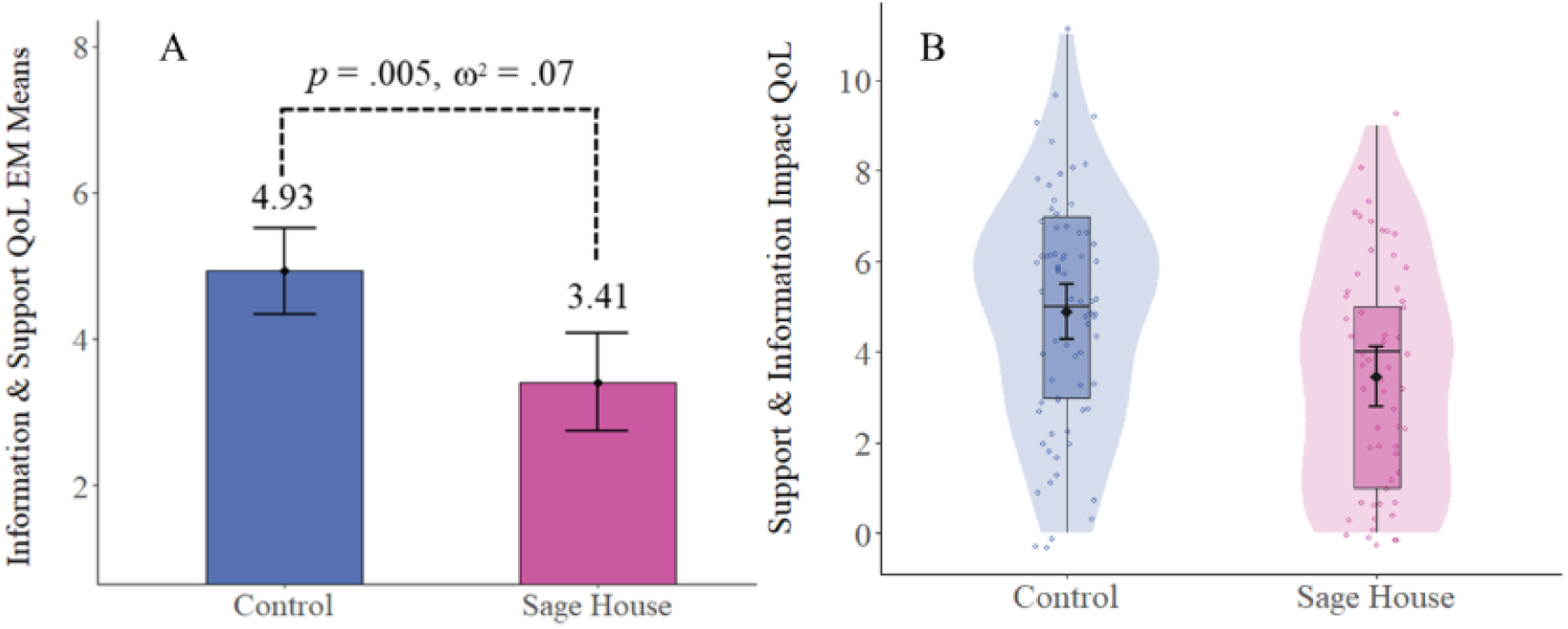
Access to Support and Information Subscale SIDECAR group comparison. A) Estimated marginal means and 95% CI after correcting for FAI. FDR corrected P-Value displayed. B) Right hand violin graphs represent raw means, 95% CI and the data distribution.

To add context the relationship between caregiver burden and the support and information subscale was examined for each experimental group. Firstly, the association was examined in the control group finding a significant positive correlation with higher impact scores being associated with higher caregiver burden, *r*(70) = .31, 95% CI [.08, .51], *p* = .008. However, when the relationship was examined in the group that had access to Sage House it did not reach the threshold to be considered statistically significant, *r*(55) = .13, 95% CI [-.12, .39], *p* = .307.

## Discussion

### Overview of Results

In the current paper we examined the research question: does access to Sage House influence key wellbeing outcomes for people with dementia and their care partners?

Compared to usual care, individuals with dementia who accessed Sage House reported higher QoL, life satisfaction, and subjective wellbeing. Therefore, the group with access was observed to score favourably on several key metrics that relate to the concept of “living well”. For care partners, while no direct impact on general wellbeing was found, the group that had access to Sage House reported significantly greater Needs-Based QoL indicating improved access to support and information. When considering this observation, it suggests that although supportive care may not be able to eliminate the challenges faced by care partners it could be a useful avenue to mitigate barriers such as poor access to information and support. Taken together, these results highlight the potential of the Sage House Model to support both people living with dementia and their care partners.

### The Results in Context

The observed wellbeing benefits may plausibly arise from key integrated care mechanisms embedded within the Sage House Model, including co-location of service, continuity of relational support, and reduced navigation burden for families. With each of these factors being highlighted as important in a series of interviews conducted with people living with dementia and care partners currently attending the centre (King et al., in preparation). When compared against existing models, the Sage House Model stands out due to its collaborative partnership structure, which integrates a broader range of local community-based services into a single location. This allows it to complement clinical services and could potentially provide a more sustainable route for implementing long term personalised multicomponent interventions within the local community, aligning with current governmental plans (Department of Health and Social Care, 2025).

Supporting people with dementia and care partners to “live well” is a priority in both UK and global governmental strategies (DoH, 2009, 2016; WHO, 2017). Therefore, the Sage House Model’s potential to enhance wellbeing for people with dementia, whilst simultaneously supporting their care partners is highly relevant. Of pertinence in the present study individuals with dementia who had access to the Sage House centre in Tangmere scored higher on all three components of the living well construct, offering a promising pathway to achieve governmental goals (Clare et al., 2014; Martyr et al., 2018; Quinn et al., 2022). Moreover, our results align with previous literature that demonstrated that multicomponent supportive care approaches can influence a range of important wellbeing and functional outcomes (Ballard et al., 2018, 2020; Eloniemi-Sulkava et al., 2009; Guzzon et al., 2023; Steinbeisser et al., 2020).

The scope of the Sage House Model was outlined in the introduction in relation to the model’s capacity to deliver services aligned with the core elements of dementia support identified by Bamford et al. (2021). This framework has been proposed as a benchmark to guide commissioners in identifying appropriate and effective post diagnostic care (Bamford et al., 2023). Therefore, within this context, the breadth of the Sage House Model combined with the observation that those with access have greater wellbeing is particularly noteworthy and underscores the need for further evaluation and larger scale studies focusing on the consideration of scalability.

### Methodological Considerations

The wellbeing measures utilised in the current study were validated and demonstrated as appropriate for the target population. However, the use of standard questionnaires to assess wellbeing brings some limitations, with participants directly commented on unrelated factors that had impacted their responses (e.g. medical problems) and how certain questions may not capture the impact of the intervention. Overall, this meant it was hard to isolate the impact of the integrated support intervention. Further work would benefit from using a wellbeing measure that can be tailored specifically to the intervention or from adopting a qualitative methodology that enable a more nuanced examination exploring how the different components of the intervention can contribute to improving wellbeing.

Although the personalised nature of how people access Sage House is a strength of the model it does provide important limitation within a research context due to the inability to standardise engagement with the different components of the model. For example, in the current study some care partners solely accessed Daybreaks for respite, whereas others engaged in support groups, activity, and training. It is recommended that future studies systematically capture patterns of service use to support more targeted research questions (e.g. do carer support groups improve wellbeing?) and enable appropriate control for variables such as frequency of access. Further work collecting this type of data is essential to understand how the type and intensity of engagement with a multicomponent supportive care approach, such as Sage House, can influence wellbeing outcomes.

The study employed a natural experiment to compare outcomes for individuals accessing the Sage House centre with those receiving standard care. Although randomised controlled trials offer strong internal validity, randomisation was not ethically appropriate in this context, given the provision of an established community service. Therefore, the use of a natural experiment following the GRACE principles (Dreyer et al., 2010, 2016) provided a pragmatic and ethically appropriate approach to evaluate the model under routine service conditions. While self-selection into the Sage House Model cannot be fully ruled out, adherence to these principles supports transparent and rigorous interpretation of the findings with real-world relevance.

It is also important to consider the broad definition of “standard care” which incorporated access to standard medical care, in addition to accessing any local services that were available to the participants (non-integrated care). Based on qualitative feedback received in the open question incorporated into this study (reported elsewhere) it was evident that our standard care control group had very disparate experiences of accessing local support services. Moving forward it is recommended that more detailed information regarding what services have been accessed should be collected to help to fully define “standard care”. Ultimately, a more direct comparison against specific models of dementia support would be optimal. It is noted that given the broad reach of the JDR network this is a possibility for future studies that have greater scope and duration.

When utilising the JDR network it should be acknowledged that the increased reach may be accompanied by some degree of sample bias. In the current study the participants recruited through the JDR network were more highly educated and less cognitively impaired. Future work using the JDR network should aim to match groups on demographics and clinical characteristics. Another sample-based consideration is that our current sample is predominantly White British, underscoring the need for research with a more diverse sample to explore whether the Sage House Model can meet varying cultural needs and contexts. Although broadly representative of the Tangmere area, as replication of the model takes place in other locations, adherence to best-practice recommendations for inclusive recruitment and reporting will be important in future work to enhance equity and representation (Brijnath et al., 2022).

### Future Directions

Future large-scale research can build on the current study by adopting a mixed factorial design, comparing individuals with access to Sage House to a more precisely defined standard care control group, and following them from diagnosis through to end of life. This approach would support the examination of broader outcomes, such as the maintenance of cognition and functional independence over time, in addition to opening the opportunity to examine important metrics such as frailty and expansion of the consideration of cost-effectiveness (e.g., employment, delayed institutionalisation).

## Conclusion

Findings from this study suggest that the Sage House Model offers a promising approach to improving key wellbeing outcomes for people with dementia (QoL, life satisfaction and subjective wellbeing) and enhancing access to support and information for care partners. The study therefore contributes to the integrated care literature by providing empirical evidence that a community-based, third sector-led integrated care model can improve wellbeing outcomes for both people living with dementia and their care partners in real-world settings. Although evaluated within the UK context, the Sage House Model reflects principles of integrated community-based dementia care that are transferable across health and social care systems internationally. For commissioners and service planners, these findings indicate that investment in community-based integrated dementia hubs may offer a feasible non-pharmacological approach to addressing post-diagnostic gaps and improving wellbeing. The Sage House Model further demonstrates how third-sector infrastructure can complement statutory services to deliver integrated post-diagnostic support at scale.

## Data Availability

All data produced in the present study are available upon reasonable request to the authors

## Acknowledgements

The authors would like to thank Sally Tabbner and the Sage House team for their collaborative and supportive approach. We would also like to extend our heartfelt gratitude to the participants who kindly gave their time to enable the project.

## References

Agüero-Torres, H., Fratiglioni, L., Guo, Z., Viitanen, M., Von Strauss, E., & Winblad, B. (1998). Dementia Is the Major Cause of Functional Dependence in the Elderly: 3-Year Follow-up Data From a Population-Based Study. American Journal of Public Health, 88(10). 10.2105/ajph.88.10.1452

Alzheimer’s Society. (2024). How many people have dementia in the UK? https://www.Alzheimers.Org.Uk/Blog/How-Many-People-Have-Dementia-Uk.

Arblaster, K., & Brennen, S. (2022). Left to Cope Alone. https://www.alzheimers.org.uk/about-us/policy-and-influencing/left-cope-alone-unmet-support-needs-after-dementia-diagnosis#

Ballard, C., Corbett, A., Orrell, M., Williams, G., Moniz-Cook, E., Romeo, R., Woods, B., Garrod, L., Testad, I., Woodward-Carlton, B., Wenborn, J., Knapp, M., & Fossey, J. (2018). Impact of person-centred care training and person-centred activities on quality of life, agitation, and antipsychotic use in people with dementia living in nursing homes: A cluster-randomised controlled trial. PLoS Medicine, 15(2). 10.1371/journal.pmed.1002500

Ballard, C., Orrell, M., Moniz-Cook, E., Woods, R., Whitaker, R., Corbett, A., Aarsland, D., Murray, J., Lawrence, V., Testad, I., Knapp, M., Romeo, R., Zala, D., Stafford, J., Hoare, Z., Garrod, L., Sun, Y., McLaughlin, E., Woodward-Carlton, B., … Fossey, J. (2020). Improving mental health and reducing antipsychotic use in people with dementia in care homes: The WHELD research programme including two RCTs. Programme Grants for Applied Research, 8. 10.3310/pgfar08060

Bamford, C., Wheatley, A., Brunskill, G., Booi, L., Allan, L., Banerjee, S., Dening, K. H., Manthorpe, J., & Robinson, L. (2021). Key components of post-diagnostic support for people with dementia and their carers: A qualitative study. PLoS ONE, 16(12 December). 10.1371/journal.pone.0260506

Bamford, C., Wilcock, J., Brunskill, G., Wheatley, A., Dening, K. H., Manthorpe, J., Allan, L., Banerjee, S., Booi, L., Griffiths, S., Rait, G., Walters, K., & Robinson, L. (2023). Improving primary care based post-diagnostic support for people living with dementia and carers: Developing a complex intervention using the Theory of Change. PLoS ONE, 18(5 May), 1–21. 10.1371/journal.pone.0283818

Banerjee, S., Smith, S. C., Lamping, D. L., Harwood, R. H., Foley, B., Smith, P., Murray, J., Prince, M., Levin, E., Mann, A., & Knapp, M. (2006). Quality of life in dementia: More than just cognition. An analysis of associations with quality of life in dementia. Journal of Neurology, Neurosurgery and Psychiatry, 77(2), 146–148. 10.1136/jnnp.2005.072983

Benjamini, Y., & Hochberg, Y. (1995). Controlling the False Discovery Rate: A Practical and Powerful Approach to Multiple Testing. The Journal of the Royal Statistical Society, Series B (Statistical Methodology), 61(1), 1–15. 10.1111/j.2517-6161.1995.tb02031.x

Ben-Shachar, M., Lüdecke, D., & Makowski, D. (2020). effectsize: Estimation of Effect Size Indices and Standardized Parameters. Journal of Open Source Software, 5(56), 2815. 10.21105/joss.02815

Brijnath, B., Croy, S., Sabates, J., Thodis, A., Ellis, S., de Crespigny, F., Moxey, A., Day, R., Dobson, A., Elliott, C., Etherington, C., Geronimo, M. A., Hlis, D., Lampit, A., Low, L. F., Straiton, N., & Temple, J. (2022). Including ethnic minorities in dementia research: Recommendations from a scoping review. In Alzheimer’s and Dementia: Translational Research and Clinical Interventions (Vol. 8, Issue 1). John Wiley and Sons Inc. 10.1002/trc2.12222

Chen, Y., Bandosz, P., Stoye, G., Liu, Y., Wu, Y., Lobanov-Rostovsky, S., French, E., Kivimaki, M., Livingston, G., Liao, J., & Brunner, E. J. (2023). Dementia incidence trend in England and Wales, 2002–19, and projection for dementia burden to 2040: analysis of data from the English Longitudinal Study of Ageing. The Lancet Public Health, 8(11), e859–e867. 10.1016/S2468-2667(23)00214-1

Clare, L., Nelis, S. M., Quinn, C., Martyr, A., Henderson, C., Hindle, J. V., Jones, I. R., Jones, R. W., Knapp, M., Kopelman, M. D., Morris, R. G., Pickett, J. A., Rusted, J. M., Savitch, N. M., Thom, J. M., & Victor, C. R. (2014). Improving the experience of dementia and enhancing active life - living well with dementia: Study protocol for the IDEAL study. Health and Quality of Life Outcomes, 12(1). 10.1186/s12955-014-0164-6

Clare, L., Wu, Y.-T., Quinn, C., Jones, I. R., Victor, C. R., Nelis, S. M., Martyr, A., Litherland, R., Pickett, J. A., Hindle, J. V, Jones, R. W., Knapp, M., Kopelman, M. D., Morris, R. G., Rusted, J. M., Thom, J. M., Lamont, R. A., Henderson, C., Rippon, I., … Behalf, O. (2019). A Comprehensive Model of Factors Associated With Capability to ‘Live Well’ for Family Caregivers of People Living With Mild-to-Moderate Dementia Findings From the IDEAL Study. http://psychology.exeter.ac.uk/reach/

Clarke, C., Woods, B., Moniz-Cook, E., Mountain, G., Øksnebjerg, L., Chattat, R., Diaz, A., Gove, D., Vernooij-Dassen, M., & Wolverson, E. (2020). Measuring the well-being of people with dementia: A conceptual scoping review. Health and Quality of Life Outcomes, 18(1), 1–14. 10.1186/s12955-020-01440-x

Dautzenberg, G., Lijmer, J., & Beekman, A. (2020). Diagnostic accuracy of the Montreal Cognitive Assessment (MoCA) for cognitive screening in old age psychiatry: Determining cutoff scores in clinical practice. Avoiding spectrum bias caused by healthy controls. International Journal of Geriatric Psychiatry, 35(3), 261–269. 10.1002/gps.5227

Department of Health. (2009). Living Well With Dementia: A National Dementia Strategy. In Living Well with Dementia A National Dementia Strategy (Vol. 2009, Issue 5). https://www.gov.uk/government/uploads/system/uploads/attachment_data/file/168220/dh_094051.pdf

Department of Health. (2016). Prime Minister’s Challenge on Dementia 2020 - Implementation Plan (Issue March 2016). https://www.gov.uk/government/uploads/system/uploads/attachment_data/file/507981/PM_Dementia-main_acc.pdf

Department of Health and Social Care. (2025, July 3). Fit for the future: 10-year health plan for England. https://www.gov.uk/government/publications/10-year-health-plan-for-england-fit-for-the-future

Dreyer, N. A., Bryant, A., & Velentgas, P. (2016). The GRACE checklist: A validated assessment tool for high quality observational studies of comparative effectiveness. Journal of Managed Care and Specialty Pharmacy, 22(10), 1107–1113. 10.18553/jmcp.2016.22.10.1107

Dreyer, N. A., Schneeweiss, S., McNeil, B. J., Berger, M. L., Walker, A. M., Ollendorf, D. A., & Gliklich, R. E. (2010). GRACE principles: Recognizing high-quality observational studies of comparative effectiveness. American Journal of Managed Care, 16(6), 467–471.

Eloniemi-Sulkava, U., Saarenheimo, M., Laakkonen, M. L., Pietilä, M., Savikko, N., Kautiainen, H., Tilvis, R. S., & Pitkälä, K. H. (2009). Family care as collaboration: Effectiveness of a multicomponent support program for elderly couples with dementia. Randomized controlled intervention study. Journal of the American Geriatrics Society, 57(12), 2200–2208. 10.1111/j.1532-5415.2009.02564.x

Farina, N., Page, T. E., Daley, S., Brown, A., Bowling, A., Basset, T., Livingston, G., Knapp, M., Murray, J., & Banerjee, S. (2017). Factors associated with the quality of life of family carers of people with dementia: A systematic review. Alzheimer’s and Dementia, 13(5), 572–581. 10.1016/j.jalz.2016.12.010

Gale, S. A., Acar, D., & Daffner, K. R. (2018). Dementia. In American Journal of Medicine (Vol. 131, Issue 10, pp. 1161–1169). Elsevier Inc. 10.1016/j.amjmed.2018.01.022

Giebel, C. M., Sutcliffe, C., Stolt, M., Karlsson, S., Renom-Guiteras, A., Soto, M., Verbeek, H., Zabalegui, A., & Challis, D. (2014). Deterioration of basic activities of daily living and their impact on quality of life across different cognitive stages of dementia: A European study. International Psychogeriatrics, 26(8), 1283–1293. 10.1017/S1041610214000775

Gilsenan, J., Gorman, C., & Shevlin, M. (2023). Explaining caregiver burden in a large sample of UK dementia caregivers: The role of contextual factors, behavioural problems, psychological resilience, and anticipatory grief. Aging and Mental Health, 27(7), 1274–1281. 10.1080/13607863.2022.2102138

Guzzon, A., Rebba, V., Paccagnella, O., Rigon, M., & Boniolo, G. (2023). The value of supportive care: A systematic review of cost-effectiveness of nonpharmacological interventions for dementia. In PLoS ONE (Vol. 18, Issue 5 May). 10.1371/journal.pone.0285305

Hargreaves, S. & Sbaffi, L. (2022). Information seeking amongst informal caregivers of people with dementia: a qualitative study. Journal of Documentation, 79(2), 281–300.

Heun, R., Burkart, M., Maier, W., & Bech, P. (1999). Internal and external validity of the WHO Well-Being Scale in the elderly general population. Acta Psychiatrica Scandinavica, 99(3), 171–178. 10.1111/j.1600-0447.1999.tb00973.x

Horton, M. C., Oyebode, J., Clare, L., Megson, M., Shearsmith, L., Brayne, C., Kind, P., Hoare, Z., Al Janabi, H., Hewison, V., Tennant, A., & Wright, P. (2021). Measuring Quality of Life in Carers of People with Dementia: Development and Psychometric Evaluation of Scales measuring the Impact of DEmentia on CARers (SIDECAR). Gerontologist, 61(3), E1–E11. 10.1093/geront/gnz136

Institute of Medicine. (2015). Living well with chronic illness: a call for public health action. Military Medicine, 180(5), 485–487. 10.7205/MILMED-D-15-00034

Lamont, R. A., Nelis, S. M., Quinn, C., Martyr, A., Rippon, I., Kopelman, M. D., Hindle, J. V., Jones, R. W., Litherland, R., & Clare, L. (2020). Psychological predictors of ‘living well’ with dementia: findings from the IDEAL study. Aging and Mental Health, 24(6), 956–964. 10.1080/13607863.2019.1566811

Lenth, R. (2023). emmeans: Estimated Marginal Means, aka Least-Squares Means_. R package version 1.8.9.

Logsdon, R. G., Gibbons, L. E. S. M. M., & Teri, L. (2002). Assessing Quality of Life in Older Adults With Cognitive Impairment. Psychosomatic Medicine, 64(3), 510–519. 10.1097/00006842-200205000-00016.

MacLeod, A., Tatangelo, G., McCabe, M., & You, E. (2017). There isn’t an easy way of finding the help that’s available. Barriers and facilitators of service use among dementia family caregivers: A qualitative study. International Psychogeriatrics, 29(5), 765–776. 10.1017/S1041610216002532

Maheswaran, H., Weich, S., Powell, J., & Stewart-Brown, S. (2012). Evaluating the responsiveness of the Warwick Edinburgh Mental Well-Being Scale (WEMWBS): Group and individual level analysis. Health and Quality of Life Outcomes, 10. 10.1186/1477-7525-10-156

Martyr, A., Nelis, S. M., Quinn, C., Rusted, J. M., Morris, R. G., & Clare, L. (2019). The relationship between perceived functional difficulties and the ability to live well with mild-to-moderate dementia: Findings from the IDEAL programme. International Journal of Geriatric Psychiatry, 34(8), 1251–1261. 10.1002/gps.5128

Martyr, A., Nelis, S. M., Quinn, C., Wu, Y. T., Lamont, R. A., Henderson, C., Clarke, R., Hindle, J. V., Thom, J. M., Jones, I. R., Morris, R. G., Rusted, J. M., Victor, C. R., & Clare, L. (2018). Living well with dementia: A systematic review and correlational meta-analysis of factors associated with quality of life, well-being and life satisfaction in people with dementia. Psychological Medicine, 48(13), 2130–2139. 10.1017/S0033291718000405

Marvardi, M., Mattioli, P., Spazzafumo, L., Mastriforti, R., Rinaldi, P., Cristina Polidori, M., Cherubini, A., Quartesan, R., Bartorelli, L., Bonaiuto, S., Cucinotta, D., Di Iorio, A., Gallucci, M., Giordano, M., Martorelli, M., Masaraki, G., Nieddu, A., Pettenati, C., Putzu, P., … Mecocci, P. (2005). The Caregiver Burden Inventory in evaluating the burden of caregivers of elderly demented patients: results from a multicenter study. In Aging Clin Exp Res (Vol. 17, Issue 1).

Müller, K. (2020). here: A Simpler Way to Find Your Files_. R package version 1.0.1.

Nakayama, N., Suzuki, M., Endo, A., Nitanda, Y., Tanabe, N., Watanabe, A., Fukuda, M., & Endo, T. (2017). Impact of dementia on behavioral independence and disturbance. Geriatrics and Gerontology International, 17(4), 605–613. 10.1111/ggi.12767

Nasreddine, Z. S., Phillips, N. A., Bédirian, V., Charbonneau, S., Whitehead, V., Collin, I., Cummings, J. L., & Chertkow, H. (2005). The Montreal Cognitive Assessment, MoCA: A brief screening tool for mild cognitive impairment. Journal of the American Geriatrics Society, 53(4), 695–699. 10.1111/j.1532-5415.2005.53221.x

NHS England. (2016). NHS England Transformation Framework – the well pathway for dementia. https://www.england.nhs.uk/mentalhealth/wp-content/uploads/sites/29/2016/03/dementia-well-pathway.pdf

Novak, M., & Guest, C. (1989). Application of a Multidimensional Caregiver Burden Inventory. The Gerontologist, 29(6), 798–803. 10.1093/geront/29.6.798

Oyebode, J. R., Pini, S., Ingleson, E., Megson, M., Horton, M., Clare, L., Al-Janabi, H., Brayne, C., & Wright, P. (2018). Development of an Item Pool for a Needs-Based Measure of Quality of Life of Carers of a Family Member with Dementia. Patient, 12(1), 125–136. 10.1007/s40271-018-0334-4

Pavot, W., & Diener, E. (1993). Review of the Satisfaction With Life Scale. Journal of Personality Assessment, 5(2), 164–172. 10.1207/s15327752jpa4901

Pavot, W., & Diener, E. (2008). The Satisfaction With Life Scale and the emerging construct of life satisfaction. Journal of Positive Psychology, 3(2), 137–152. 10.1080/17439760701756946

Pavot, W., Diener, E., & Colvin, C. R. (1991). Further Validation of the Satisfaction With Life Scale: Evidence for the Cross-Method Convergence of Well-Being Measures. Journal of Personality Assessment, 57(1), 149–161. 10.1207/s15327752jpa5701_17

Peterson, K., Hahn, H., Lee, A. J., Madison, C. A., & Atri, A. (2016). In the Information Age, do dementia caregivers get the information they need? Semi-structured interviews to determine informal caregivers’ education needs, barriers, and preferences. BMC Geriatrics, 16(1). 10.1186/s12877-016-0338-7

Quinn, C., Nelis, S. M., Martyr, A., Morris, R. G., Victor, C., & Clare, L. (2020). Caregiver influences on ‘living well’ for people with dementia: Findings from the IDEAL study. Aging and Mental Health, 24(9), 1505–1513. 10.1080/13607863.2019.1602590

Quinn, C., Pickett, J. A., Litherland, R., Morris, R. G., Martyr, A., & Clare, L. (2022). Living well with dementia: What is possible and how to promote it. International Journal of Geriatric Psychiatry, 37(1), 1–7. 10.1002/gps.5627

R Core Team. (2023). R: A Language and Environment for Statistical Computing. R Foundation for Statistical Computing.

Reid, B., & O’Brien, L. (2021). The psychological effects of caring for a family member with dementia. Nursing Older People, 33(6), 21–27. 10.7748/nop.2021.e1295

Revelle, W. (2023). psych: Procedures for Psychological, Psychometric, and Personality Research. Northwestern University.

Schuling, J., de Haan, R., Limburg, M., & Groenier, K. H. (1993). The Frenchay Activities Index: Assessment of functional status in stroke patients. Stroke, 24(8), 1173–1177. 10.1161/01.STR.24.8.1173

Segal, M. E., & Schall, R. R. (1994). Determining Functional/Health Status and Its Relation to Disability in Stroke Survivors. http://ahajournals.org

Steinbeisser, K., Schwarzkopf, L., Graessel, E., & Seidl, H. (2020). Cost-effectiveness of a non-pharmacological treatment vs. “care as usual” in day care centers for community-dwelling older people with cognitive impairment: results from the German randomized controlled DeTaMAKS-trial. European Journal of Health Economics, 21(6), 825–844. 10.1007/s10198-020-01175-y

Tennant, R., Hiller, L., Fishwick, R., Platt, S., Joseph, S., Weich, S., Parkinson, J., Secker, J., & Stewart-Brown, S. (2007). The Warwick-Dinburgh mental well-being scale (WEMWBS): Development and UK validation. Health and Quality of Life Outcomes, 5. 10.1186/1477-7525-5-63

Thorgrimsen, L. Bs. S. A. Mrcp. S. A. P. R. L. Ms. de M. L. M. Ms. W. R. T. Ms. O. M. PhD. W. Q. of L. I. I. A. (2003). The Validity and Reliability of the Quality of Life-Alzheimer’s Disease (QoL-AD) Scale. Alzheimer Disease & Associated Disorders, 17(4), 201–208.

Topp, C. W., Østergaard, S. D., Søndergaard, S., & Bech, P. (2015). The WHO-5 well-being index: A systematic review of the literature. Psychotherapy and Psychosomatics, 84(3), 167–176. 10.1159/000376585

Tournier, I., Orton, L., Dening, T., Ahmed, A., Holthoff-Detto, V., & Niedderer, K. (2023). An Investigation of the Wishes, Needs, Opportunities and Challenges of Accessing Meaningful Activities for People Living with Mild to Moderate Dementia. International Journal of Environmental Research and Public Health, 20(7). 10.3390/ijerph20075358

van Horik, J. O., Collins, R., Martyr, A., Henderson, C., Jones, R. W., Knapp, M., Quinn, C., Thom, J. M., Victor, C., & Clare, L. (2022). Limited receipt of support services among people with mild-to-moderate dementia: Findings from the IDEAL cohort. International Journal of Geriatric Psychiatry, 37(3), 10–11. 10.1002/gps.5688

Visentin, D. C., Cleary, M., & Hunt, G. E. (2020). The earnestness of being important: Reporting non-significant statistical results. In Journal of Advanced Nursing (Vol. 76, Issue 4, pp. 917–919). Blackwell Publishing Ltd. 10.1111/jan.14283

World Health Organization. (2025, March 31). Dementia. https://www.who.int/news-room/fact-sheets/detail/dementia

Wickham, H., Averick, M., Bryan, J., Chang, W., MMcGowan, L. D., François, R., Grolemund, G., Hayes, A., Henry, L., Hester, J., Kuhn, M., Pedersen, T. L., Miller, E., Bache, S. M., Müller, K., Ooms, J., Robinson, D., Seidel, D. P., Spinu, V., … Woo, H. (2019). Welcome to the tidyverse. Journal of Open Source Software, 4(43), 1–6.

World Health Organization. (1998). Wellbeing Measures in Primary Health Care: The DEPCARE Project. WHO Regional Office for Europe. https://iris.who.int/bitstream/handle/10665/349766/WHO-EURO-1998-4234-43993-62027-eng.pdf?sequence=1&isAllowed=y

World Health Organization. (2017). Global action plan on the public health response to dementia 2017 - 2025. Geneva: World Health Organization. https://www.who.int/publications/i/item/global-action-plan-on-the-public-health-response-to-dementia-2017---2025

Zhang, X., Astivia, O. L. O., Kroc, E., & Zumbo, B. D. (2022). How to Think Clearly About the Central Limit Theorem. Psychological Methods, 28(6), 1427–1445. 10.1037/met0000448

