## Supplementary Information for "A Community-Based Integrated Partnership Approach to Living Well with Dementia: Evaluating the Impact of the Sage House Model"

### Supplementary Materials

#### Supplementary Table 1

*Sample Characteristics for Participants with Dementia*

| **Measure** | **People with Dementia** | | | |
|---|---|---|---|---|
| | **Combined Sample** | **Control** | **Sage House** | **Sig** |
| ***N*** | 135 | 89 | 46 | |
| **Age** | 74.64 (SD 8.3) | 72.25 (8.12) | 79.26 (6.56) | < .001 |
| **Time Since Diagnosis** | 3 years (SD 2.9) | 3.37 (3.11) | 2.48 (2.23) | *.151* |
| **Frenchay Activities Index** | 26.61 (9.45) | 28.99 (8.14) | 22.02 (10.17) | < .001 |
| **MoCa** (*n* = 127: 87/40) | 18.24 (6.53) | 19.70 (6.30) | 15.05 (5.91) | < .001 |
| **Gender** | | | | |
| Male | 75 (56%) | 50 (56%) | 25 (54%) | .807 |
| Female | 58 (43%) | 37 (42%) | 21 (46%) | |
| Non-Binary | 2(2%) | 2 (2%) | - | |
| **Education Level** | | | | |
| Secondary or below | 29 (22%) | 15 (17%) | 14 (30%) | .046 |
| Post-Secondary | 37 (27%) | 19 (21%) | 18 (39%) | |
| University Degree | 35 (26%) | 25 (28%) | 10 (22%) | |
| University Post Grad + | 30 (22%) | 26 (29%) | 4 (9%) | |
| Unknown | 4 (3%) | 4 (5%) | - | |
| **Dementia Type** | | | | |
| Dementia - unknown | 24 (18%) | 8 (9%) | 16 (35%) | |
| Mild Cognitive Impairment | 20 (15%) | 17 (19%) | 3 (7%) | |
| Alzheimer’s Disease | 51(38%) | 30 (34%) | 21 (46%) | |
| Early Onset Dementia | 8 (6%) | 7 (8%) | 1 (2%) | |
| Frontotemporal Dementia | 3 (2%) | 3 (3%) | - | |
| Lew Body Dementia | 5 (4%) | 5 (6%) | - | |
| Vascular Dementia | 13 (10%) | 9 (10%) | 4 (9%) | |
| Mixed Dementia | 11 (10%) | 10 (11%) | 1 (2%) | |
| **Ethnicity/ Nationality** | | | | |
| White British | 112 (83%) | 69 (78%) | 43 (94%) | |
| White Irish | 5 (4%) | 4 (5%) | 1 (2%) | |
| White Welsh | 2 (1.5%) | 2 (2%) | - | |
| White Scottish | - | - | - | |
| White Other | 11 (8%) | 11 (12%) | - | |
| Asian British | 2 (1.5%) | 2 (2%) | - | |
| Cornish | 1 (0.74%) | 1 (1%) | - | |
| Canadian | 1 (0.74%) | - | 1 (2%) | |
| Black British/Caribbean | - | - | - | |
| Unknown | 1 (0.74%) | - | 1 (2%) | |

Abbreviations: FAI, Frenchay Activities Index. Sig, Significance
Note: For continuous data Kruskal Wallis test was utilised and for frequency data Fisher’s exact test

### Supplementary Materials

#### Supplementary Table 2
*Sample Characteristics for Care Partner Participants*

| **Measure** | **Care Partners** | | | |
|---|---|---|---|---|
| | **Combined** | **Control** | **Sage House** | **Sig** |
| ***N*** | 129 | 72 | 57 | |
| **Age** | 67.23 (SD 9.84) | 65.40 (SD 9.88) | 69.54 (SD 9.38) | .016 |
| **Proxy FAI** | 15.29 (SD 8.96) | 16.35 (SD 9.43) | 13.97 (SD 8.23) | .143 |
| **Care Giver Burden** | 41.19 (16.02) | 38.53 (SD 15.54) | 44.54 (SD 16.11) | .038 |
| **Gender** | | | | |
| Male | 97 (75%) | 14 (19%) | 18 (32%) | .151 |
| Female | 32 (25%) | 58 (81%) | 39 (68%) | |
| Non-Binary | - | - | - | |
| **Education Level** | | | | |
| Secondary or below | 21 (16%) | 7 (10%) | 14 (25%) | .010 |
| Post-Secondary | 43 (33%) | 19 (26%) | 24 (42%) | |
| University Degree | 37 (29%) | 25 (35%) | 12 (21%) | |
| University Post Grad + | 28 (22%) | 17 (24%) | 6 (10%) | |
| Unknown | 5 (4%) | 4 (5%) | 1 (2%) | |
| **Ethnicity/ Nationality** | | | | |
| White British | 118 (92%)) | 62 (86%) | 56 (98%) | |
| White Irish | - | - | - | |
| White Welsh | 1 (0.78%) | 1 (1%) | | |
| White Scottish | 1 (0.78%) | 1 (1%) | | |
| White Other | 4 (3%) | 3 (4%) | 1 (2%) | |
| Asian British | 2 (1.5%) | 2 (2%) | | |
| Cornish | - | - | - | |
| Canadian | - | - | | |
| Black British/Caribbean | 1 (0.78%) | 1 (1%)) | | |
| Unknown | 2 (1.5%) | 2 (3%) | | |

Abbreviations: FAI, Frenchay Activities Index. Sig, Significance
Note: For continuous data Kruskal Wallis test was utilised and for frequency data Fisher's exact test

**Supplementary Materials**

**Supplementary Table 3**
*GRACE Checklist Mapping*

| GRACE Principle | Application in This Study |
|---|---|
| Concurrent comparator | Both intervention and comparison groups were recruited during the same period and assessed by the same team. |
| Adjustment for confounds | ANCOVA used to control for differences in functional independence. |
| Outcome consistency | Protocols were applied uniformly across groups, with standardisation training provided to research staff. |
| Objective, standardised outcome | Standardised, validated wellbeing measures with established psychometric properties were used. |
| Sensitivity Analysis | No formal sensitivity analysis was performed; however, results were examined with and without any significant outliers to confirm removal had no impact on the statistical conclusions drawn (robustness checking). |
| Follow up duration sufficient | Participants in the intervention group had accessed the Centre for a minimum of one month prior to outcome assessment, allowing sufficient exposure to service to assess impact. |

The research was conducted under routine practice conditions, against a realistic standard care comparator, sources of bias have been acknowledged (e.g., natural experiment, JDR recruitment etc.), clinically meaningful outcomes have been utilized (e.g., living well construct) and the model (due to its community-based co-design approach) should be applicable to both UK and internal settings.
